# Melanocortin-1 receptor (*MC1R*) genotypes do not correlate with size in two cohorts of medium-to-giant congenital melanocytic nevi

**DOI:** 10.1101/2020.04.10.20055301

**Authors:** Neus Calbet-Llopart, Mirella Pascini-Garrigos, Gemma Tell-Martí, Miriam Potrony, Vanessa Martins da Silva, Alicia Barreiro, Susana Puig, Guillaume Captier, Isabelle James, Nathalie Degardin, Cristina Carrera, Josep Malvehy, Heather C. Etchevers, Joan Anton Puig-Butillé

**Affiliations:** Dermatology Department, Melanoma Unit, Hospital Clínic de Barcelona, IDIBAPS, University of Barcelona, Barcelona, Spain; Centro de Investigación Biomédica en Red de Enfermedades Raras (CIBERER), Instituto de Salud Carlos III, Barcelona, Spain.; Plastic pediatric surgery, University of Montpellier Hospital, Montpellier, France; Service de Chirurgie Réparatrice de l’Enfant, Clinique du Val d’Ouest, Ecully, France; Service de Chirurgie Plastique Réparatrice, Hôpital de la Timone Enfants, Marseille, France; Aix-Marseille Univ, Marseille Medical Genetics, INSERM, U1251, Faculté de Médecine, Marseille, France; Molecular Biology CORE, Biomedical Diagnostic Center (CDB), Melanoma Unit, Hospital Clínic de Barcelona, IDIBAPS, University of Barcelona, Spain

**Keywords:** Congenital melanocytic nevi or nevus, melanocortin 1 receptor (MC1R) gene, nevogenesis, cutaneous lesion, inherited variants, Giant Congenital Melanocytic Nevus (GCMN), phenotype, population

## Abstract

Congenital melanocytic nevi (CMN) are cutaneous malformations whose prevalence is inversely correlated with projected adult size. CMN are caused by somatic mutations, but epidemiological studies suggest that germline genetic factors may influence CMN development. In CMN patients from the U.K., genetic variants in the *MC1R* gene, such as p.V92M and loss-of-function variants, have been previously associated with larger CMN. We analyzed the association of *MC1R* variants with CMN characteristics in 113 medium-to-giant CMN patients from Spain and from a distinct cohort of 53 patients from France, Norway, Canada and the U.S. These cohorts were similar at the clinical and phenotypical level, except for the number of nevi per patient. We found that the p.V92M or loss-of-function *MC1R* variants either alone or in combination did not correlate with CMN size, in contrast to the U.K. CMN patients. An additional case-control analysis with 259 unaffected Spanish individuals, showed a higher frequency of *MC1R* compound heterozygous or homozygous variant genotypes in Spanish CMN patients compared to the control population (15.9% vs. 9.3%; *P*=0.075). Altogether, this study suggests that *MC1R* variants are not associated with CMN size in these non-U.K. cohorts. Additional studies are required to define the potential role of *MC1R* as a risk factor in CMN development.

**SIGNIFICANCE:** Congenital melanocytic nevi (CMN) are common pigmented lesions that originate during prenatal life, without clear evidence of a genetic predisposition. To date, limited data exist regarding the role of the *MC1R* gene, a key regulator of human pigmentation, in the development of the class of rarer CMN that are greater than 10 cm diameter at projected adult size and associated with increased morbidity or mortality risks. This study provides data from a large set of such CMN patients to support the hypothesis that MC1R could be involved in the development of these types of lesions, but at the same time discounting its influence on the size of CMN across distinct populations. Improving our understanding of genetic susceptibility to rare types of CMN is necessary to determine whether routine germline genotyping is relevant in clinical practice.

## 1. INTRODUCTION

Congenital melanocytic nevi (CMN) are benign melanocytic tumors of the skin, which are present at birth or become visibly pigmented during the first years of life (Price & Schaffer, 2010). CMN are classified based on the projected adult size (PAS) of the largest lesion (Krengel, Scope, Dusza, Vonthein, & Marghoob, 2013). Large (PAS 20-40 cm in diameter) and giant (PAS > 40 cm) CMN are rare lesions found in approximately 1/20,000 to 1/50,000-100,000 newborns, respectively (Alikhan, Ibrahimi, & Eisen, 2012). This subset of patients has an increased risk of developing pediatric and adult melanoma within the lesion, the viscera or the central nervous system (CNS) (Kinsler et al., 2017). Furthermore, these patients may present other CNS abnormalities, including neurocutaneous melanosis and brain tumors (Foster et al., 2001; Jakchairoongruang, Khakoo, Beckwith, & Barkovich, 2018), Dandy-Walker malformations (De Cock, Snauwaert, Van Rompaey, Morren, & Demaerel, 2014; Marnet et al., 2009; Schreml et al., 2008; Walbert, Sloan, Cohen, & Koubeissi, 2009), arachnoid cysts (Peters, Jansen, & Engelbrecht, 2000), tethered spinal cord (Foster et al., 2001; Tian, Foster, Jakacki, Reyes-Mugica, & Greene, 2015), hydrocephalus (Hsueh, Ho, Chiu, & Shen, 2004; Peters et al., 2000) or epilepsy (Wen et al., 2001). Large and giant CMN may occur in isolation or as part of a syndrome with variable phenotypic expression (Kinsler, Shaw, Merks, & Hennekam, 2012a). CMN lesions are characterized according to a consensus classification including the anatomic location, further broken down into stereotypical distribution patterns (the “6B” scheme” in giant CMN (Martins da Silva et al., 2017) and the “biker glove” pattern (Kittler, Mathes, Kinsler, & Frieden, 2019); color heterogeneity; surface rugosity; presence of hypertrichosis; nodularity; and numbers of “satellite” or multiple CMN that appear during the first years of life or are visible at birth (Krengel et al., 2013). In this paper, we use the term “multiple CMN” instead of “satellite” and in contrast to “single CMN”, when the patient exhibits more than one CMN with different sizes (Kinsler, 2011).

CMN seem to be caused by the acquisition of a postzygotic somatic mutation that constitutively activates the MAPK signaling pathway in a melanocyte-competent cell lineage. These events include oncogenic point mutations in the *BRAF* and *NRAS* genes, which are the most recurrent alterations (Bauer, Curtin, Pinkel, & Bastian, 2007; Charbel et al., 2014; Polubothu et al., 2019), but also chromosomal rearrangements (Baltres et al., 2019; Dessars et al., 2007; Martins da Silva et al., 2019). Although CMN are the result of somatic mosaicism, epidemiological data and case reports of familial recurrence in up to 25% of second-degree relatives, as opposed to approximately 1% of the general population, suggest the existence of a germline predisposition for CMN (Danarti, Konig, & Happle, 2003; de Wijn, Zaal, Hennekam, & van der Horst, 2010; Kinsler, Birley, & Atherton, 2009).

A study conducted in CMN patients from the U.K. concluded that the presence of germline variants in the melanocortin-1-receptor (*MC1R*) gene was both a risk factor for CMN development and that the presence of certain germline variants may modulate the size of the CMN (Kinsler et al., 2012b). The *MC1R* gene, a key regulator of human pigmentation (Dessinioti, Antoniou, Katsambas, & Stratigos, 2011; Sturm, 2009), is highly polymorphic in populations of European origin (Gerstenblith, Goldstein, Fargnoli, Peris, & Landi, 2007). Many *MC1R* variants are hypomorphic alleles that cause various degrees of loss of the receptor’s ability to activate eumelanin synthesis. Some of these are strongly associated with the “red hair color” (RHC) phenotype, characterized by fair skin, red hair, freckles, high UV radiation sensitivity and lack of tanning ability (Valverde, Healy, Jackson, Rees & Thody, 1995). The most common RHC variants have been classified according to their phenotypic penetrance into high-penetrance “R” or lower-penetrance “r” alleles (reviewed in Herraiz, Garcia-Borron, Jiménez-Cervantes & Olivares, 2017). Both R and r alleles have also been associated with increased melanoma risk (Hu et al., 2014; Palmer et al., 2000; Raimondi et al., 2008; Tagliabue et al., 2018; Williams, Olsen, Hayward, & Whiteman, 2011) or with a specific melanoma clinicopathological subtype (Puig-Butille et al., 2013), particularly in R/R but also R/r combinations. These “red hair” alleles also exert similar influence on melanoma risk, hair color and skin phototype in Mediterranean populations, such as the Spanish (Fernandez et al., 2007). Indeed, most but not all people with red hair do carry two *MC1R* variants, some of whom express “r” variants in epistasis with other gene loci, yet people carrying two variant *MC1R* alleles are more likely to have light brown or blonde hair than red (Morgan et al., 2018). Within the U.K. CMN cohort, *MC1R* status, comprising a p.V92M “r” allele, any “R” allele, or both, when taken together, was significantly associated with increasing CMN size, most so for those patients with G2-sized giant CMN (PAS > 60 cm) (Kinsler et al., 2012b). This study aims to analyze the impact of *MC1R* variants on phenotypic attributes of CMN in two multicentric cohorts of medium-to-giant CMN patients from different European and North American countries.

## 2. MATERIALS AND METHODS

### 2.1. Participants

Each study participant and/or their parent or legal guardian signed written informed consent, as appropriate. All aspects of this study comply with the declaration of Helsinki.

Overall, the study included 166 patients with medium, large or giant CMN from independent fair-skinned populations. In all cases, CMN lesions were phenotypically classified following the latest consensus classification (Krengel et al., 2013) (**Table 1**), and giant CMN were additionally classified following the 6B guidelines (Martins da Silva et al., 2017) (**Supporting table S1**).

**Table 1.** Clinical features and evaluation of the CMN phenotypic characteristics of the Spanish and Marseille CMN patient cohorts

| Patients' clinical features |  | All CMN patients<br>(N=166) | Spanish cohort<br>(N=113) | Marseille cohort<br>(N=53) | P-value<br>(Spanish vs. Marseille) |
| --- | --- | --- | --- | --- | --- |
| **Age in years** (mean ± SD) | | 16.81 ± 16.54 | 17.13 ± 16.65 | 16.14 ± 16.44 | 0.720 |
| **Sex** | Male | 38.0% (62) | 41.6% (47) | 30.0% (15) | 0.160 |
|  | Female | 62.0% (101) | 58.4% (66) | 70.0% (35) |  |
|  | Missing | 3 | 0 | 3 |  |
| **Hair Color** | Red | 2.2% (3) | 1.0% (1) | 6.3% (2) | 0.154 |
|  | Blond | 20.0% (27) | 17.5% (18) | 28.1% (9) |  |
|  | Brown | 67.4% (91) | 70.9% (73) | 56.3% (18) |  |
|  | Black | 10.4% (14) | 10.7% (11) | 9.4% (3) |  |
|  | Missing | 31 | 10 | 21 |  |
| CMN phenotypic features |  | All CMN patients<br>(N=166) | Spanish cohort<br>(N=113) | Marseille cohort<br>(N=53) | P-value<br>(Spanish vs. Marseille) |
| **Size (PAS)** | Medium | 24.7% (41) | 27.4% (31) | 18.9% (10) | 0.121 |
|  | Large | 16.9% (28) | 19.5% (22) | 11.3% (6) |  |
|  | Giant | 58.4% (97) | 53.1% (60) | 69.8% (37) |  |
| **Anatomic location** | Only head | 21.2% (35) | 23.9% (27) | 15.4% (8) | 0.462 |
|  | Including trunk | 66.1% (109) | 63.7% (72) | 71.2% (37) |  |
|  | Only extremities | 12.7% (21) | 12.4% (14) | 13.5% (7) |  |
|  | Missing | 1 | 0 | 1 |  |
| **Color heterogeneity** | None | 35.3% (54) | 30.9% (34) | 46.5% (20) | 0.141 |
|  | Moderate | 43.1% (66) | 44.5% (49) | 39.5% (17) |  |
|  | Marked | 21.6% (33) | 24.5% (27) | 14.0% (6) |  |
|  | Missing | 13 | 3 | 10 |  |
| **Multiple CMN count** | 0 | 28.0% (46) | 35.1% (39) | 13.2% (7) | **0.002** |
|  | <20 | 26.8% (44) | 28.8% (32) | 22.6% (12) |  |
|  | 20-50 | 12.8% (21) | 12.6% (14) | 13.2% (7) |  |
|  | >50 | 32.3% (53) | 23.4% (26) | 50.9% (27) |  |
|  | Missing | 2 | 2 | 0 |  |
| **Surface rugosity** | None | 49.7% (76) | 44.6% (50) | 63.4% (26) | 0.062 |
|  | Moderate | 41.2% (63) | 43.8% (49) | 34.1% (14) |  |
|  | Marked | 9.2% (14) | 11.6% (13) | 2.4% (1) |  |
|  | Missing | 13 | 1 | 12 |  |
| **Nodules** | None | 70.9% (107) | 74.1% (83) | 61.5% (24) | 0.282 |
|  | Scattered | 17.9% (27) | 15.2% (17) | 25.6% (10) |  |
|  | Extensive | 11.3% (17) | 10.7% (12) | 12.8% (5) |  |
|  | Missing | 15 | 1 | 14 |  |
| **Hypertrichosis** | None | 21.0% (30) | 17.9% (20) | 32.3% (10) | 0.116 |
|  | Notable | 50.3% (72) | 54.5% (61) | 35.5% (11) |  |
|  | Marked | 28.7% (41) | 27.7% (31) | 32.3% (10) |  |
|  | Missing | 23 | 1 | 22 |  |

Abbreviation: CMN, congenital melanocytic nevi.

#### 2.1.1. Spanish CMN Patient Cohort

The Spanish cohort included 113 patients from Spain recruited at the Hospital Clínic of Barcelona (HCB) and through the Spanish association of patients with large or giant CMN (Asociación Española de Nevus Gigante Congénito or Asonevus). Clinical and phenotypic data were obtained by direct examination and/or digital photographs by trained dermatologists from the HCB patients and by self-reported questionnaires from the Asonevus patients. The Asonevus patients were encouraged to answer the questionnaire with the guidance of their dermatologist or pediatrician, and to attach photos and reports of neonatologists, pediatricians, dermatologists and plastic surgeons they might have consulted, in order to assess the accuracy of the phenotypic reporting. Adequate blood or saliva samples for DNA extraction were obtained from HCB patients. The Asonevus patients received a saliva collection kit with the corresponding instructions for sample collection alongside the questionnaire.

This study was approved by the Clinical Research Ethics Committee of the HCB.

#### 2.1.2. Marseille CMN Patient Cohort

The Marseille cohort included 53 patients from different populations that were recruited through a multicentric study based at the Aix-Marseille University: three from Norway, 11 from France, two from Canada and 37 from the United States. Only phototypes I to IV were included in this study in accordance with the composition of the Spanish cohort. The parents of the French pediatric patients completed phenotyping questionnaires with their referring plastic surgeons. Photographs were provided in addition to nevus and non-nevus (unaffected skin, blood or saliva) samples. The other CMN patients were recruited at the 2010 Nevus Outreach International Conference, completing a similar questionnaire in the presence of a pediatric dermatologist and providing blood samples.

This study was approved by the ethical committee CPP Sud-Méditerranée II (214-C03 from 11 April 2014) and received the French Ministry of Research authorization (DC2013-1769).

#### 2.1.3. Spanish control cohort

In addition to medium-to-giant CMN patients, we included a set of 259 individuals as a control population in order to obtain the allelic frequency of *MC1R* variants in the Spanish population. The control individuals were adults (≥18 years old) considered healthy (able to perform normal activities and in the case of any chronic condition, this was treated and under control), with none of the following criteria: (i) personal history of melanoma, non-cutaneous malignancies, immunosuppression, or genodermatosis predisposing to skin cancer (i.e., xeroderma pigmentosum, albinism, or Gorlin syndrome), (ii) familial history of melanoma in first-degree relatives, (iii) pregnant women, and (iv) relatives of another control individual in the same study.

### 2.2. *MC1R* Genotyping

Genomic DNA was isolated from peripheral blood lymphocytes or from epithelial cells in saliva samples. Genomic DNA from the Spanish cohort blood samples was isolated using the CMG-715 Chemagic™ DNA blood kit with the automated method Chemagic™ MSM1 (Chemagen, Baesweiler, Germany), or using an Autopure LS (Qiagen, Hilden, Germany) workflow from the Marseille cohort samples. Saliva was collected in OG-500 or OG-575 Oragene® saliva collection kits, depending on the age of the patient, and DNA was extracted using the prepIT®-L2P reagents (DNAGenotek, Ontario, Canada). Polymerase chain reaction (PCR) was used to amplify two overlapping fragments of the *MC1R* coding region using the following primers: NT-F, 5’-TGTAAAACGACGGCCAGTGCAGCACCATGAACTAAGCA-3’ together with TM-R, 5’-CAGGAAACAGCTATGACCTTTAAGGCCAAAGCCCTGGT-3’; and CT-R, 5’-CAGGAAACAGCTATGACCCAGGGTCACACAGGAACCA-3’ together with TM-F, 5’-TGTAAAACGACGGCCAGTAACCTGCACTCACCCATGTA-3’. The thermal cycling conditions were as follows: 1 cycle of denaturation at 95°C for 5 min, 35 amplification cycles (94°C for 1 min, 55°C for 1 min, and 72°C for 3 min), and a final extension at 72°C for 10 min. The entire *MC1R* coding region was sequenced using universal M13 primers by GENEWIZ (Takeley, UK). Sequences were analyzed using SeqPilot 4.0.1 software (JSI Medical Systems, Ettenheim, Germany).

### 2.3. Statistical Analysis

*MC1R* non-synonymous variants were classified as high-penetrance “R” or low-penetrance “r” alleles according to previously reported criteria (García-Borrón, Sánchez-Laorden & Jiménez-Cervantes, 2005; Kinsler et al., 2012b; Raimondi et al., 2008; Vallone et al., 2018). *MC1R* variants classified as “R” were p.D84E, p.R142H, p.R151C, p.I155T, p.R160W, p.R163*, and p.D294H. All other non-synonymous variants, including p.V60L, p.V92M, and p.R163Q, were classified as “r”. Synonymous variants were considered equivalent to wild-type *MC1R* alleles. Statistical analysis was performed using the IBM® SPSS® Statistics software package version 20 (IBM Corp., Armonk, N.Y., USA). Pearson’s chi-squared and Student’s t-tests were used to compare categorical and continuous variables, respectively. All tests were two-sided and considered statistically significant if the p-value was <0.05.

### 2.4. Data Availability

Datasets related to this study are available upon request from the corresponding author at Hospital Clinic of Barcelona.

## 3. RESULTS

### 3.1. Clinical and Phenotypical Characteristics of CMN Patients

Two cohorts of medium-to-giant CMN patients were ascertained; these were designated as the Spanish Cohort (113 CMN patients recruited at Hospital Clínic of Barcelona or through the Spanish association of patients with large or giant CMN (Asonevus) and the Marseille Cohort (53 CMN patients from France, Norway, Canada, and the United States recruited through a multicentric study based at the Aix-Marseille University). Comparison of clinical and phenotypical features between the Spanish and the Marseille cohorts showed no statistical differences in terms of patient age, sex, hair color and projected adult size (PAS) or anatomic locations of the lesions (**Table 1**).

We categorized all lesions according to the latest consensus classification (Krengel et al., 2013) (**Table 1**). The subset of giant CMN lesions (N=97), which accounted for 53.1% and 69.8% of the Spanish and the Marseille cohorts, respectively, were also classified according to 6B body distribution patterns (Martins da Silva et al., 2017) (**Supporting table S1**). Both cohorts were similar at the clinical and phenotypic level except for the number of multiple CMN in the patient (*P*=0.002) (**Table 1**). Patients with >50 multiple CMN were nearly twice as frequent in the Marseille cohort compared to the Spanish cohort. This difference between cohorts was restricted to the subset of CMN patients classified as giant (**Supporting table S2**).

### 3.2. Molecular Screening of *MC1R* variants

We detected nine recurrent non-synonymous *MC1R* variants in CMN patients, including the “R” variants p.D84E, p.R142H, p.R151C, p.I155T, p.R160W, p.D294H, and “r” variants p.V60L, p.V92M, p.R163Q (**Table 2**). In addition, six uncommon *MC1R* non-synonymous variants were detected in seven patients (p.R163*, p.A81P, p.S83P, p.R142C, p.V122M, and p.T262S). The Spanish and the Marseille cohort showed significant differences in the allelic frequency of the p.V92M variant (*P*=0.002) and, to a lesser extent, the p.D294H variant (*P*=0.023) (**Table 2**). Differences in the allelic frequency of p.V92M were restricted to the subset of patients with CMN classified as giant (N=97) (**Supporting table S3**). Overall, we found non-synonymous *MC1R* variants in 63.9% of CMN patients, corresponding to 59.3% and 73.6% of the Spanish and the Marseille cohorts, respectively. Although we found no statistically significant differences between both cohorts in terms of the prevalence of *MC1R* genotypes, we observed a lower overall fraction of *MC1R* variant carriers and compound heterozygous or homozygous *MC1R* genotypes among the Spanish patients (**Table 3**).

**Table 2.** Allelic frequency of the most common non-synonymous *MC1R* variants in CMN patient cohorts

| *MC1R* gene |  |  | Minor allele frequencies |  |  |  |
| --- | --- | --- | --- | --- | --- | --- |
| Amino Acid change | Alleles | Minor allele | All CMN patients (N=166) | Spanish cohort (N=113) | Marseille cohort (N=53) | P-value (Spanish vs. Marseille) |
| **p.V60L<sup>†</sup>** | G/T | T | 0.17 | 0.18 | 0.13 | 0.260 |
| **p.V92M<sup>†</sup>** | G/A | A | 0.07 | 0.04 | 0.13 | **0.002** |
| **p.R163Q<sup>†</sup>** | G/A | A | 0.04 | 0.03 | 0.06 | 0.717 |
| **p.D84E<sup>‡</sup>** | C/A | A | 0.02 | 0.01 | 0.03 | 0.175 |
| **p.R142H<sup>‡</sup>** | G/A | A | 0.02 | 0.02 | 0.00 | 0.123 |
| **p.R151C<sup>‡</sup>** | C/T | T | 0.04 | 0.03 | 0.06 | 0.262 |
| **p.I155T<sup>‡</sup>** | T/C | C | 0.02 | 0.02 | 0.02 | 0.941 |
| **p.R160W<sup>‡</sup>** | C/T | T | 0.02 | 0.02 | 0.02 | 0.941 |
| **p.D294H<sup>‡</sup>** | G/C | C | 0.02 | 0.01 | 0.05 | **0.023** |

<sup>†</sup> Low-penetrance RHC variants (“r” variants)
<sup>‡</sup> High-penetrance RHC variants (“R” variants)
Abbreviation: CMN, congenital melanocytic nevi; RHC, red hair color.

**Table 3.**
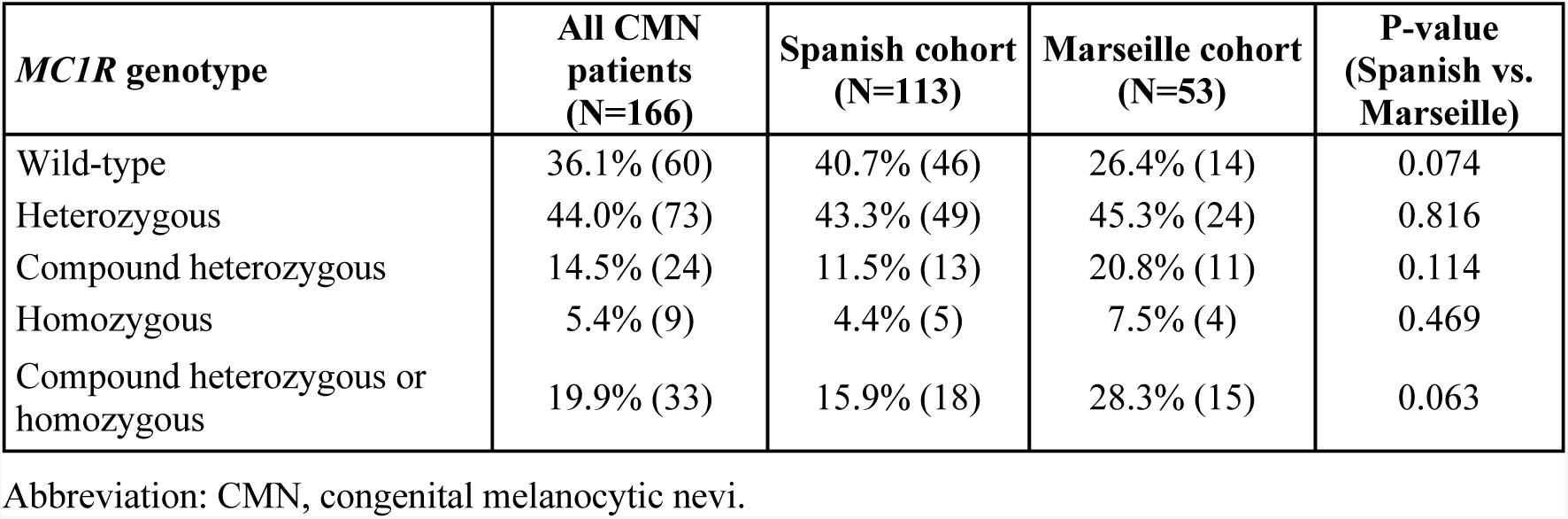
Prevalence of heterozygous or homozygous non-synonymous *MC1R* variants in CMN patients

First, we evaluated whether the presence of *MC1R* variants had an impact on the phenotypic features of CMN. We did not observe any significant association between the presence of *MC1R* variants and PAS or anatomic location of the lesion. For instance, giant CMN patients with the same characteristics in terms of multiple CMN count and *MC1R* genotype showed different clinical presentations of the CMN (**Figure 1**).

**Figure 1.**
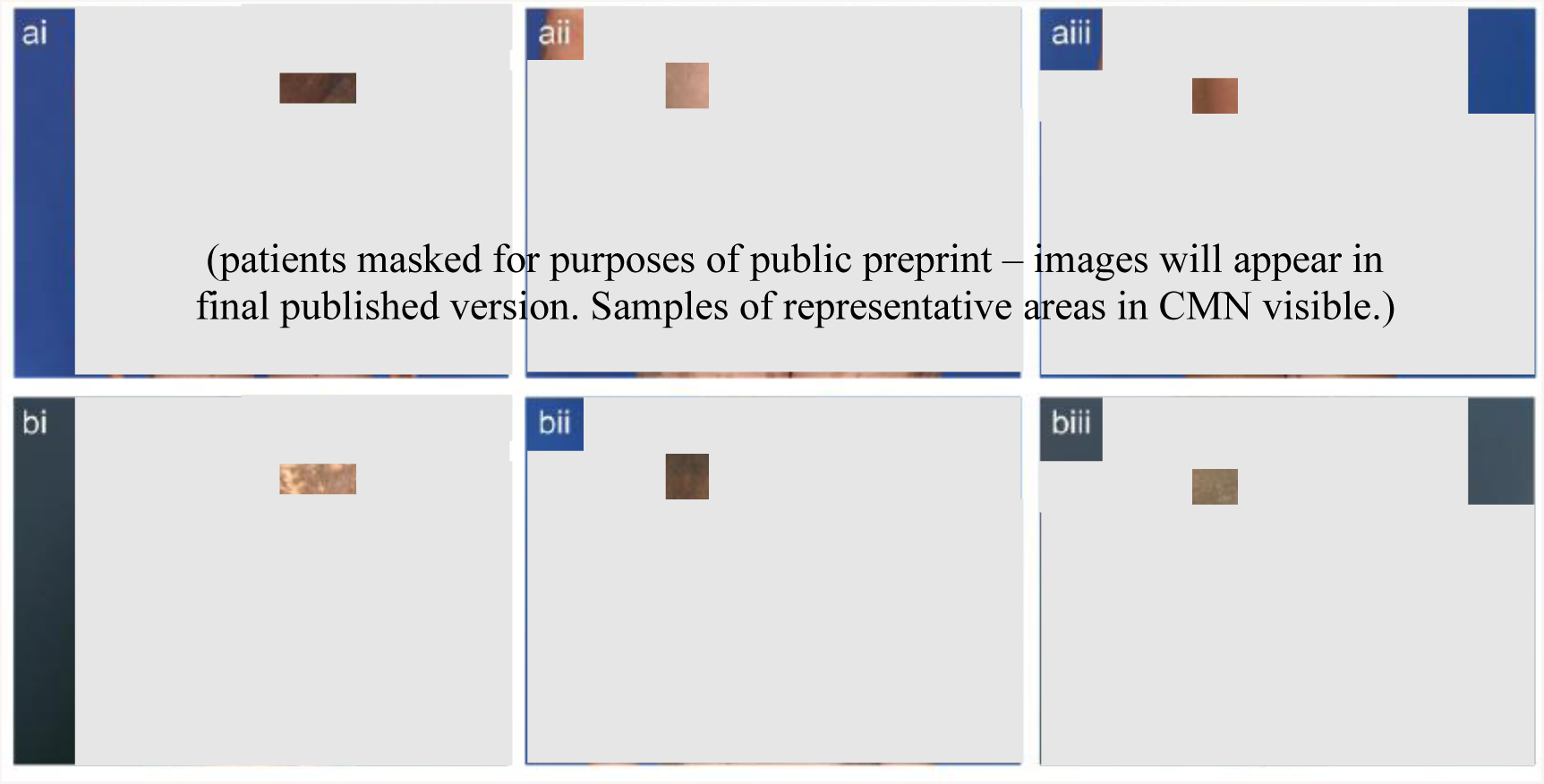
Correlation of *MC1R* genotype and phenotypic features of patients with giant congenital melanocytic nevi (CMN). Examples of whole-body photography of female patients with CMN classified as G2 with >50 multiple CMN with either **(ai-iii)** bathing trunk distribution or **(bi-iii)** bolero distribution, with different *MC1R* genotypes and different CMN presentations. **(i)** Wild-type *MC1R*, **(ii)** presence of one *MC1R* variant (p.V60L), and **(iii)** presence of two *MC1R* variants (**(a)** p.R151C, p.R163Q; **(b)** p.V60L, p.R151C). Written, informed consent was obtained for image publication in all cases.

Based on the previous findings observed in U.K. CMN patients (Kinsler et al., 2012b), we specifically evaluated the association of the p.V92M variant or “R” *MC1R* alleles with PAS of the lesions in our CMN patients (**Table 4**). We did not observe any association between the p.V92M variant and PAS of the CMN when all patients were analyzed together. The presence of any “R” *MC1R* variant alone or in combination with the p.V92M variant was also not associated with CMN size differences. However, when we analyzed each cohort separately, we found in the Spanish cohort that the allelic frequency of the p.V92M variant was lower in CMN patients with higher PAS (*P*=0.025). The p.V92M variant was observed in 12.9% and 18.2% of Spanish medium and large CMN patients, respectively, but only in 1.7% of Spanish giant CMN patients. Giant CMN patients differed in the number of multiple CMN between cohorts (**Supporting table S2**). Thus, we assessed whether the number of multiple CMN was a confounding factor for the association between the p.V92M variant and the size of the lesion observed in Spanish patients (**Supporting Table S4**). We did not find a significant association between the number of multiple CMN and the presence of the p.V92M variant in either CMN cohort, indicating that these were unrelated variables.

**Table 4.** Association of the *MC1R* genotype with projected adult size (PAS) of the CMN.

| Cohort | CMN size | *MC1R* genotype |  |  |  |  |  |  |  |  |
| --- | --- | --- | --- | --- | --- | --- | --- | --- | --- | --- |
|  |  | Presence of p.V92M variant |  |  | Presence of “R” variant |  |  | Presence of p.V92M or “R” variant |  |  |
|  |  | No | Yes | P-value | No | Yes | P-value | No | Yes | P-value |
| **All CMN patients**<br>N=166 | Medium | 82.9% (34) | 17.1% (7) | 0.175 | 82.9% (34) | 17.1% (7) | 0.183 | 65.9% (27) | 34.1% (14) | 0.976 |
|  | Large | 78.6% (22) | 21.4% (6) |  | 82.1% (23) | 17.9% (5) |  | 64.3% (18) | 35.7% (10) |  |
|  | Giant | 90.7% (88) | 9.3% (9) |  | 70.1% (68) | 29.9% (29) |  | 63.9% (62) | 36.1% (35) |  |
| **Spanish cohort**<br>N=113 | Medium | 87.1% (27) | 12.9% (4) | **0.025** | 87.1% (27) | 12.9% (4) | 0.382 | 74.2% (23) | 25.8% (8) | 0.820 |
|  | Large | 81.8% (18) | 18.2% (4) |  | 81.8% (18) | 18.2% (4) |  | 68.2% (15) | 31.8% (7) |  |
|  | Giant | 98.3% (59) | 1.7% (1) |  | 75.0% (45) | 25.0% (15) |  | 75.0% (45) | 25.0% (15) |  |
| **Marseille cohort**<br>N=53 | Medium | 70.0% (7) | 30.0% (3) | 0.748 | 70.0% (7) | 30.0% (3) | 0.572 | 40.0% (4) | 60.0% (6) | 0.917 |
|  | Large | 66.7% (4) | 33.3% (2) |  | 83.3% (5) | 16.7% (1) |  | 50.0% (3) | 50.0% (3) |  |
|  | Giant | 78.4% (29) | 21.6% (8) |  | 62.2% (23) | 37.8% (14) |  | 45.9% (17) | 54.1% (20) |  |

Abbreviation: CMN, congenital melanocytic nevi; PAS, projected adult size.

CMN patients were more likely to have two *MC1R* variants (compound heterozygous or homozygous) than the U.K. control population, regardless of the particular *MC1R* variant (Kinsler et al., 2012b). To explore the potential role of the *MC1R* gene as a risk factor for CMN development, we performed a case-control analysis comparing the Spanish CMN patient cohort with 259 Spanish control individuals. Although no statistically significant differences were observed in the prevalence of *MC1R* variants between groups, we found a higher, but not statistically significant, frequency of compound heterozygous or homozygous genotypes in CMN patients compared to the control population (15.9% vs. 9.3%; *P*=0.075) (**Table 5**). In contrast, the allelic frequency of the p.V92M variant (*P*=0.868) or the presence of any “R” allele (*P*=0.815) was similar between CMN patients and control individuals (**Supporting table S5**).

**Table 5.**
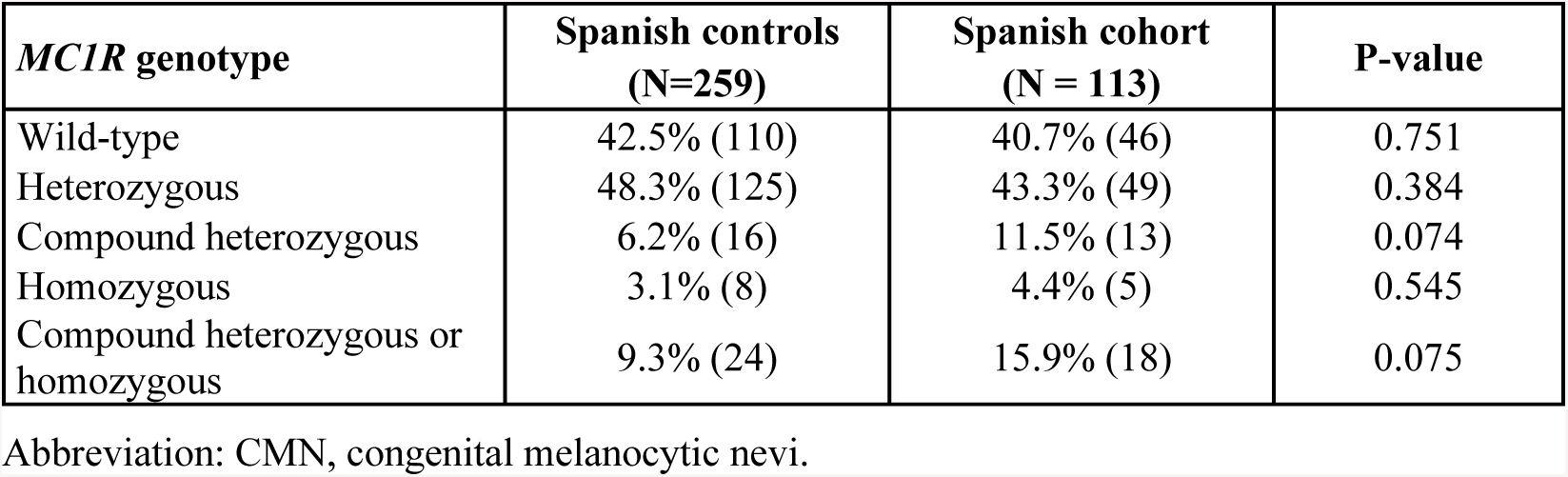
Comparison of the prevalence of heterozygous or homozygous non-synonymous *MC1R* variants between Spanish controls and Spanish CMN patients

| ***MC1R* genotype** | **Spanish controls**<br>**(N=259)** | **Spanish cohort**<br>**(N = 113)** | **P-value** |
| --- | --- | --- | --- |
| Wild-type | 42.5% (110) | 40.7% (46) | 0.751 |
| Heterozygous | 48.3% (125) | 43.3% (49) | 0.384 |
| Compound heterozygous | 6.2% (16) | 11.5% (13) | 0.074 |
| Homozygous | 3.1% (8) | 4.4% (5) | 0.545 |
| Compound heterozygous or<br>homozygous | 9.3% (24) | 15.9% (18) | 0.075 |

Abbreviation: CMN, congenital melanocytic nevi.

## 4. DISCUSSION

In the present study, we have constituted and compared two previously unpublished independent cohorts of medium-to-giant CMN patients from different populations: one exclusively from Spain (Spanish cohort) and another from diverse origins recruited at Aix-Marseille University (Marseille cohort). We analyzed all CMN patients together, as these cohorts exhibit similar clinical and phenotypic features, except for the number of multiple CMN that may accompany the principal lesion (“satellites”). Giant CMN are significantly associated with higher numbers of such multiple CMN and a higher prevalence of other clinical signs of CMN syndrome, such as melanoma or neurocutaneous melanosis (Marghoob, Dusza, Oliveria, & Halpern, 2004; Martins da Silva et al., 2017; Price et al., 2015). In the present study, the fraction of giant CMN patients was greater in the Marseille cohort (69.8%) compared to the Spanish cohort (53.1%). However, the difference in the number of multiple CMN was restricted to the subset of giant CMN patients, suggesting that molecular differences may exist between these subsets.

Based on a previously published cohort of CMN patients from the U. K. (Kinsler et al., 2012b), we assessed the role of certain *MC1R* variants in CMN development. We analyzed nearly twice as many CMN patients as compared to the U. K. cohort (N=166 vs. N=84) and with a higher representation of giant CMN (58% vs. 49%). In contrast to the earlier findings, in our study the presence of the p.V92M variant and/or any “R” alleles, either alone or in combination, was not associated with a larger PAS of the CMN. By analyzing the cohorts independently, we found that the presence of the p.V92M variant even protected against developing the largest CMN in the Spanish cohort.

The allelic frequency of *MC1R* variants, including the p.V92M variant, differs among populations, being lower in Mediterranean populations compared with northern European populations (Dessinioti et al., 2011; Gerstenblith et al., 2007). Likewise, the Spanish CMN patients carried fewer *MC1R* variants and had a lower frequency of the p.V92M variant compared with the Marseille cohort, which had more diverse origins. In contrast, the prevalence of *MC1R* genotypes was similar between the Marseille and the U. K. cohorts (Kinsler et al., 2012b). In the U. K. cohort, 52% of giant CMN patients carried either an “R” allele or the p.V92M variant, very similar to the Marseille cohort, where 54% of giant CMN patients carried these variants but where the effect of *MC1R* on size was not replicated. A major drawback of studying such a rare and heterogeneous condition is that it is likely that subdivision into regions too small to be proxies for populations diminishes our capacity to distinguish real from spurious associations. Thus, the significant association of the p.V92M variant with a lower PAS of the CMN in Spanish patients might be a spurious result caused by both the low frequency of the variant and the limited size of the giant CMN patient subset. Children from dark-skinned populations of sub-Saharan Africa, in whom non-synonymous *MC1R* variants are rare (reviewed in Herraiz et al., 2017), also develop the largest, G2-type CMN (Katibi, Ogunbiyi, Brown & Adeyemi, 2014; Endomba, Mbega, Tochie & Petnga, 2018). Altogether, these data indicate that *MC1R* genotype is not likely to contribute to a larger CMN sizeCMN.

In the U. K. study, the risk for CMN development was associated with the number of *MC1R* variants rather than with the presence of a particular variant (Kinsler et al., 2012b). Similarly, in our Spanish cohort, we have detected a difference in the prevalence of compound heterozygous or homozygous genotypes between patients and controls. The lower proportion of *MC1R* variant carriers in the Spanish population probably affects these results. In the future, further case-control studies should be conducted in larger populations with a high prevalence of *MC1R* variants to resolve whether the *MC1R* genotype has an impact on any relevant aspect of CMN development.

In conclusion, our study suggests that the *MC1R* genotype is not associated with the size of CMN. However, we cannot rule out the role of *MC1R* as a risk factor for CMN development, especially in carriers of *MC1R* variants on both alleles. Additional studies in other populations, including detailed clinical descriptions following the consensus classifications of all lesions and of the individuals who carry them, are necessary to clearly elucidate the role of the *MC1R* gene in CMN development.

## CONFLICT OF INTEREST

The authors state no conflict of interest.

## ACKNOWLEDGMENTS

The authors warmly thank the patient advocacy groups Asociación Española de Nevus Gigante Congénito (Asonevus; Spain), Nevus Outreach (U.S.A.), Association Nævus 2000 France-Europe and the Association du Nævus Géant Congénital (France) for both their financial support of this study and their invaluable assistance in recruiting many of its subjects. We also thank Dr. Veronica Kinsler for her assistance in phenotyping a large proportion of the patients from Nevus Outreach, Inc. We are grateful to our patients and relatives, and to physicians and nurses from the Melanoma Unit of Hospital Clínic of Barcelona, specially to Isabel del Busto MD, Maite Elisa Eliceche MD, Alexandra Franco, Abel Caño, Asunción Arnaiz and Beatriz Alejo for helping to collect patient data and samples; and to Judit Mateu from the “Melanoma: image, genetics and immunology” group at IDIBAPS for her technical assistance. The research at the Melanoma Unit from Hospital Clinic of Barcelona is partially funded by grants PI15/00716, PI15/00956 and PI18/00959 from Fondo de Investigaciones Sanitarias, Spain; the Spanish Federation of Neuromuscular Disease (FEDASEM), the Spanish Federation of Rare Diseases (FEDER), and Isabel Gemio Research Foundation of muscular dystrophy and other rare diseases (FIG) through the Call for Research Projects on Rare Diseases 2014 through the initiative “We Are Rare, All Are Unique, by the CIBER de Enfermedades Raras of the Instituto de Salud Carlos III, Spain, co-funded by ISCIII-Subdirección General de Evaluación and European Regional Development Fund (ERDF), a way to build Europe”; AGAUR 2017_SGR_1134 of the Catalan Government, Spain; by the European Commission under the 7th Framework Programme (Diagnoptics); by CERCA Programme / Generalitat de Catalunya, Spain; and by the Leo Messi Foundation. Part of the work was developed at the building Centro Esther Koplowitz, Barcelona. Neus Calbet-Llopart is the recipient of a PhD Fellowship FPU17/05453 (FPU) from Ministerio de Educación, Cultura y Deportes, Spain. Cristina Carrera is the recipient of a research personal grant from Catalan Government through the “Pla estratègic de recerca i innovació en salut (PERIS) 2018-2020”, Ref. BDNS 357800.

## SUPPORTING INFORMATION

**Supporting table S1.** Patterns of distribution of giant CMN according to the 6B rule.

| **Giant CMN**<br>**6B distribution** | **All CMN**<br>**patients**<br>**(N=97)** | **Spanish**<br>**cohort**<br>**(N=60)** | **Marseille**<br>**cohort**<br>**(N=37)** | **P-value**<br>**(Spanish vs.**<br>**Marseille)** |
| --- | --- | --- | --- | --- |
| **Bolero** | 17.6% (29) | 28.3% (17) | 32.4% (12) | 0.479 |
| **Back** | 10.3% (17) | 15.0% (9) | 21.6% (8) |  |
| **Bathing trunk** | 23.0% (38) | 41.7% (25) | 35.1% (13) |  |
| **Breast/belly** | 1.2% (2) | 3.3% (2) | 0.0% (0) |  |
| **Body extremity** | 4.8% (8) | 6.7% (4) | 10.8% (4) |  |
| **Body** | 1.2% (2) | 5.0% (2) | 0.0% (0) |  |
| **Missing** | 1 | 1 | 0 |  |

Abbreviation: CMN, congenital melanocytic nevi.

**Supporting table S2.** Evaluation of CMN phenotypic characteristics of the Spanish and Marseille CMN patient cohorts, including only (A) medium and large CMN or (B) giant CMN

| CMN phenotypic features |  | All CMN patients<br>(N=69) | Spanish medium/large CMN cohort<br>(N=53) | Marseille medium/large CMN cohort<br>(N=16) | P-value<br>(Spanish vs. Marseille) |
| --- | --- | --- | --- | --- | --- |
| **Color heterogeneity** | None | 40.0% (26) | 34.6% (18) | 61.5% (8) | 0.151 |
|  | Moderate | 46.2% (30) | 51.9% (27) | 23.1% (3) |  |
|  | Marked | 13.8% (9) | 13.5% (7) | 15.4% (2) |  |
|  | Missing | 4 | 1 | 3 |  |
| **Multiple CMN count** | 0 | 58.2% (39) | 62.7% (32) | 43.75% (7) | 0.465 |
|  | <20 | 28.4% (19) | 23.5% (12) | 43.75% (7) |  |
|  | 20-50 | 7.5% (5) | 7.8% (4) | 6.25% (1) |  |
|  | >50 | 6.0% (4) | 5.9% (3) | 6.25% (1) |  |
|  | Missing | 2 | 2 | 0 |  |
| **Surface rugosity** | None | 62.1% (41) | 56.6% (30) | 84.6% (11) | 0.155 |
|  | Moderate | 30.3% (20) | 34.0% (18) | 15.4% (2) |  |
|  | Marked | 7.6% (5) | 9.4% (5) | 0.0% (0) |  |
|  | Missing | 3 | 0 | 3 |  |
| **Nodules** | None | 87.9% (58) | 86.8% (46) | 92.3% (12) | 0.580 |
|  | Scattered | 6.1% (4) | 7.5% (4) | 0.0% (0) |  |
|  | Extensive | 6.1% (4) | 5.7% (3) | 7.7% (1) |  |
|  | Missing | 3 | 0 | 3 |  |
| **Hypertrichosis** | None | 19.0% (12) | 15.1% (8) | 40.0% (4) | 0.094 |
|  | Notable | 58.7% (37) | 64.2% (34) | 30.0% (3) |  |
|  | Marked | 22.2% (14) | 20.8% (11) | 30.0% (3) |  |
|  | Missing | 6 | 0 | 6 |  |

**B**
| CMN phenotypic features |  | All CMN patients<br>(N=97) | Spanish giant CMN cohort<br>(N=60) | Marseille giant CMN cohort<br>(N=37) | P-value<br>(Spanish vs. Marseille) |
| --- | --- | --- | --- | --- | --- |
| **Color heterogeneity** | None | 31.8% (28) | 27.6% (16) | 40.0% (12) | 0.102 |
|  | Moderate | 40.9% (36) | 37.9% (22) | 46.7% (14) |  |
|  | Marked | 27.3% (24) | 34.5% (20) | 13.3% (4) |  |
|  | Missing | 9 | 2 | 7 |  |
| **Multiple CMN count** | 0 | 7.2% (7) | 11.7% (7) | 0.0% (0) | **0.006** |
|  | <20 | 25.8% (25) | 33.3% (20) | 13.5% (5) |  |
|  | 20-50 | 16.5% (16) | 16.7% (10) | 16.2% (6) |  |
|  | >50 | 50.5% (49) | 38.3% (23) | 70.3% (26) |  |
| **Surface rugosity** | None | 40.2% (35) | 33.9% (20) | 53.6% (15) | 0.134 |
|  | Moderate | 49.4% (43) | 52.5% (31) | 42.9% (12) |  |
|  | Marked | 10.3% (9) | 13.6% (8) | 3.6% (1) |  |
|  | Missing | 10 | 1 | 9 |  |
| **Nodules** | None | 57.6% (49) | 62.7% (37) | 46.2% (12) | 0.265 |
|  | Scattered | 27.1% (23) | 22.0% (13) | 38.5% (10) |  |
|  | Extensive | 15.3% (13) | 15.3% (9) | 15.4% (4) |  |
|  | Missing | 12 | 1 | 11 |  |
| **Hypertrichosis** | None | 22.5% (18) | 20.3% (12) | 28.6% (6) | 0.713 |
|  | Notable | 43.8% (35) | 45.8% (27) | 38.1% (8) |  |
|  | Marked | 33.8% (27) | 33.9% (20) | 33.3% (7) |  |
|  | Missing | 17 | 1 | 16 |  |

Abbreviation: CMN, congenital melanocytic nevi.

**Supporting table S3.** Allelic frequency of the most common non-synonymous *MC1R* variants in the Spanish and Marseille CMN patient cohorts, including only (A) medium and large CMN or (B) giant CMN

| *MC1R* gene |  |  | Minor allele frequencies |  |  |  |
| --- | --- | --- | --- | --- | --- | --- |
| Amino Acid change | Alleles | Minor allele | All medium/large CMN patients (N=69) | Spanish medium/large CMN cohort (N=53) | Marseille medium/large CMN cohort (N=16) | P-value (Spanish vs. Marseille) |
| p.V60L<sup>†</sup> | G/T | T | 0.15 | 0.19 | 0.03 | **0.030** |
| p.V92M<sup>†</sup> | G/A | A | 0.09 | 0.08 | 0.16 | 0.170 |
| p.R163Q<sup>†</sup> | G/A | A | 0.04 | 0.04 | 0.06 | 0.547 |
| p.D84E<sup>‡</sup> | C/A | A | 0.01 | 0.01 | 0.00 | 0.581 |
| p.R142H<sup>‡</sup> | G/A | A | 0.01 | 0.02 | 0.00 | 0.434 |
| p.R151C<sup>‡</sup> | C/T | T | 0.04 | 0.03 | 0.06 | 0.364 |
| p.I155T<sup>‡</sup> | T/C | C | 0.00 | 0.00 | 0.00 | - |
| p.R160W<sup>‡</sup> | C/T | T | 0.01 | 0.02 | 0.00 | 0.434 |
| p.D294H<sup>‡</sup> | G/C | C | 0.01 | 0.00 | 0.03 | 0.068 |

**B**
| *MC1R* gene |  |  | Minor allele frequencies |  |  |  |
| --- | --- | --- | --- | --- | --- | --- |
| Amino Acid change | Alleles | Minor allele | All giant CMN patients (N=97) | Spanish giant CMN cohort (N=60) | Marseille giant CMN cohort (N=37) | P-value (Spanish vs. Marseille) |
| p.V60L<sup>†</sup> | G/T | T | 0.18 | 0.18 | 0.18 | 0.990 |
| p.V92M<sup>†</sup> | G/A | A | 0.05 | 0.01 | 0.12 | **0.001** |
| p.R163Q<sup>†</sup> | G/A | A | 0.03 | 0.02 | 0.05 | 0.144 |
| p.D84E<sup>‡</sup> | C/A | A | 0.02 | 0.01 | 0.04 | 0.299 |
| p.R142H<sup>‡</sup> | G/A | A | 0.02 | 0.03 | 0.00 | 0.170 |
| p.R151C<sup>‡</sup> | C/T | T | 0.04 | 0.03 | 0.05 | 0.481 |
| p.I155T<sup>‡</sup> | T/C | C | 0.03 | 0.03 | 0.03 | 0.805 |
| p.R160W<sup>‡</sup> | C/T | T | 0.02 | 0.02 | 0.03 | 0.622 |
| p.D294H<sup>‡</sup> | G/C | C | 0.03 | 0.02 | 0.04 | 0.612 |

<sup>†</sup> Low-penetrance RHC variants (“r” variants)
<sup>‡</sup> High-penetrance RHC variants (“R” variants)
Abbreviation: CMN, congenital melanocytic nevi; RHC, red hair color.

**Supporting table S4.** Association of the *MC1R* genotype with the number of multiple CMN per patient.

| Patients | Multiple CMN count | *MC1R* genotype |  |  |  |  |  |  |  |  |
| --- | --- | --- | --- | --- | --- | --- | --- | --- | --- | --- |
|  |  | Presence of p.V92M variant |  |  | Presence of “R” variant |  |  | Presence of p.V92M or “R” variant |  |  |
|  |  | No | Yes | P-value | No | Yes | P-value | No | Yes | P-value |
| **All CMN patients**<br>N=164 | 0 | 82.6% (38) | 17.4% (8) | 0.428 | 82.6% (38) | 17.4% (8) | 0.509 | 65.2% (30) | 34.8% (16) | 0.995 |
|  | <20 | 86.4% (38) | 13.6% (6) |  | 72.7% (32) | 27.3% (12) |  | 63.6% (28) | 36.4% (16) |  |
|  | 20-50 | 81.0% (17) | 19.0% (4) |  | 76.2% (16) | 23.8% (5) |  | 61.9% (13) | 38.1% (8) |  |
|  | >50 | 92.5% (49) | 7.5% (4) |  | 69.8% (37) | 30.2% (16) |  | 64.2% (34) | 35.8% (19) |  |
| **Spanish cohort**<br>N=111 | 0 | 84.6% (33) | 15.4% (6) | 0.156 | 82.1% (32) | 17.9% (7) | 0.960 | 66.7% (26) | 33.3% (13) | 0.734 |
|  | <20 | 93.8% (30) | 6.3% (2) |  | 78.1% (25) | 21.9% (7) |  | 75.0% (24) | 25.0% (8) |  |
|  | 20-50 | 92.9% (13) | 7.1% (1) |  | 78.6% (11) | 21.4% (3) |  | 78.6% (11) | 21.4% (3) |  |
|  | >50 | 100.0% (26) | 0.0% (0) |  | 76.9% (20) | 23.1% (6) |  | 76.9% (20) | 23.1% (6) |  |
| **Marseille cohort**<br>N=53 | 0 | 71.4% (5) | 28.6% (2) | 0.360 | 85.7% (6) | 14.3% (1) | 0.630 | 57.1% (4) | 42.9% (3) | 0.503 |
|  | <20 | 66.7% (8) | 33.3% (4) |  | 58.3% (7) | 41.7% (5) |  | 33.3% (4) | 66.7% (8) |  |
|  | 20-50 | 57.1% (4) | 42.9% (3) |  | 71.4% (5) | 28.6% (2) |  | 28.6% (2) | 71.4% (5) |  |
|  | >50 | 85.2% (23) | 14.8% (4) |  | 63.0% (17) | 37.0% (10) |  | 51.9% (14) | 48.1% (13) |  |

Abbreviation: CMN, congenital melanocytic nevi

**Supporting table S5.**
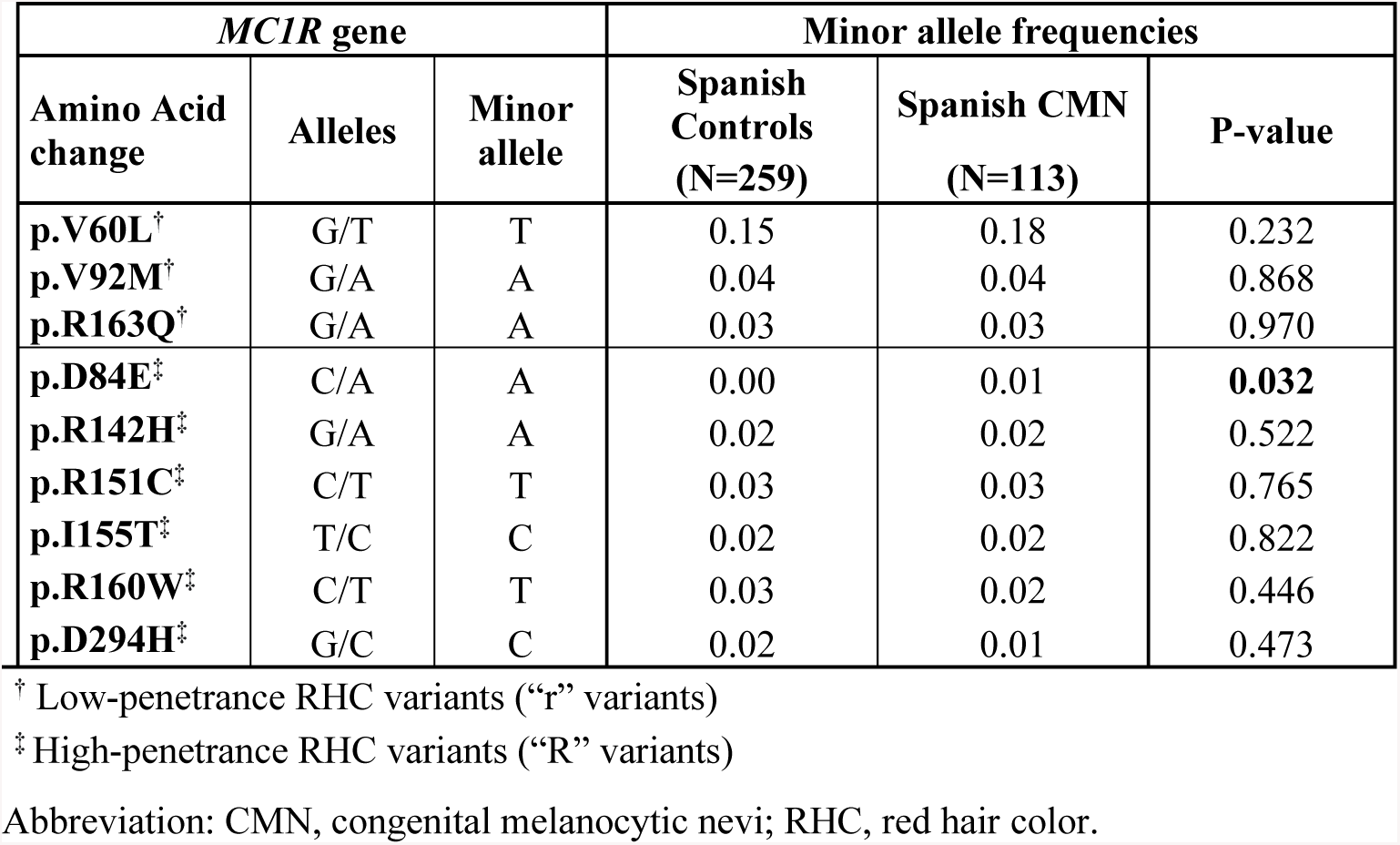
Allelic frequency of the most common non-synonymous *MC1R* variants in the Spanish controls and Spanish CMN patient cohorts

| *MC1R* gene |  |  | Minor allele frequencies |  |  |
| --- | --- | --- | --- | --- | --- |
| Amino Acid change | Alleles | Minor allele | Spanish Controls (N=259) | Spanish CMN (N=113) | P-value |
| **p.V60L<sup>†</sup>** | G/T | T | 0.15 | 0.18 | 0.232 |
| **p.V92M<sup>†</sup>** | G/A | A | 0.04 | 0.04 | 0.868 |
| **p.R163Q<sup>†</sup>** | G/A | A | 0.03 | 0.03 | 0.970 |
| **p.D84E<sup>‡</sup>** | C/A | A | 0.00 | 0.01 | **0.032** |
| **p.R142H<sup>‡</sup>** | G/A | A | 0.02 | 0.02 | 0.522 |
| **p.R151C<sup>‡</sup>** | C/T | T | 0.03 | 0.03 | 0.765 |
| **p.I155T<sup>‡</sup>** | T/C | C | 0.02 | 0.02 | 0.822 |
| **p.R160W<sup>‡</sup>** | C/T | T | 0.03 | 0.02 | 0.446 |
| **p.D294H<sup>‡</sup>** | G/C | C | 0.02 | 0.01 | 0.473 |

<sup>†</sup> Low-penetrance RHC variants (“r” variants)
<sup>‡</sup> High-penetrance RHC variants (“R” variants)
Abbreviation: CMN, congenital melanocytic nevi; RHC, red hair color.

## Notes

### Competing Interest Statement

The authors have declared no competing interest.

### Funding Statement

None of the authors or their institutions have received payment or services from a third party for any aspect of the submitted work at any time.
The authors warmly thank the patient advocacy groups Asociación Española de Nevus Gigante Congénito (Asonevus; Spain), Nevus Outreach (U.S.A.), Association Nævus 2000 France-Europe and the Association du Nævus Géant Congénital (France) for both their financial support of this study and their invaluable assistance in recruiting many of its subjects.
The research at the Melanoma Unit from Hospital Clinic of Barcelona is partially funded by grants PI15/00716, PI15/00956 and PI18/00959 from Fondo de Investigaciones Sanitarias, Spain; the Spanish Federation of Neuromuscular Disease (FEDASEM), the Spanish Federation of Rare Diseases (FEDER), and Isabel Gemio Research Foundation of muscular dystrophy and other rare diseases (FIG) through the Call for Research Projects on Rare Diseases 2014 through the initiative “We Are Rare, All Are Unique, by the CIBER de Enfermedades Raras of the Instituto de Salud Carlos III, Spain, co-funded by ISCIII-Subdirección General de Evaluación and European Regional Development Fund (ERDF), a way to build Europe”; AGAUR 2017_SGR_1134 of the Catalan Government, Spain; by the European Commission under the 7th Framework Programme (Diagnoptics); by CERCA Programme / Generalitat de Catalunya, Spain; and by the Leo Messi Foundation.
Neus Calbet-Llopart is the recipient of a PhD Fellowship FPU17/05453 (FPU) from Ministerio de Educación, Cultura y Deportes, Spain. Cristina Carrera is the recipient of a research personal grant from Catalan Government through the “Pla estratègic de recerca i innovació en salut (PERIS) 2018-2020”, Ref. BDNS 357800.

